# Indian Publications on SARS-CoV-2: A Bibliometric Study of WHO COVID-19 Database

**DOI:** 10.1101/2020.06.08.20125518

**Authors:** N. Vasantha Raju, S.B. Patil

## Abstract

Nowadays, the whole World is under threat of Coronavirus disease (COVID-19). The ongoing COVID-19 pandemic has resulted in many fatalities and forced scientific communities to foster their Research and Development (R & D) activities. As a result, there is an enormous growth of scholarly literature on the subject. In order to combat this novel coronavirus, the open access to scientific literature is essential. On this line, many reputed academic institutions and publication firms have made their literature on COVID-19 accessible to all. By maintaining the database of updated information on global literature on Coronavirus disease, the World Health Organization (WHO) is playing a pivotal role. The present study analyzed 89 Indian publications on SARS-CoV-2 accessible through WHO COVID-19 database. The research data was restricted for the period of 2/3/2020 to 12/5/2020. The analysis was carried out in light of the objectives of the study. The study found the considerable and constant growth of Indian publications on COVID-19 from mid-April. It is interesting to note that the prolific authors belong to either AIIMS or ICMR institutes. Majority of the COVID-19 articles were found to be collaborative publications. The study noticed that no research publications on COVID-19 have appeared from North Eastern region. Regarding the research output on COVID-19, the performance of largest states like Uttar Pradesh, Madhya Pradesh and Bihar was found to be poor. Delhi state contributed highest publications on COVID-19. The All India Institute of Medical Sciences (AIIMS), New Delhi was the most productive institution in terms of publications. It is also important to note that the central government undertakings like AIIMS and ICMR, New Delhi and its affiliated institutions shared largest proportion of publications on COVID-19. The Indian Journal of Medical Research has emerged as the productive journal contributing highest number of the publications. The highest contribution in COVID-19 research takes the form of journal articles. In terms of research area, the majority of the publications were related to Epidemiology. The study reported covid, coronavirus, India, pandemic, sars etc. as the frequently occurred keywords in the COVID-19 publications. The highly cited publications were of evidenced based studies. It is observed that the studies pertaining to virology, diagnosis and treatment, clinical features etc. have received highest citations than general studies on epidemiology or pandemic.

## 1. Introduction

Coronavirus or novel coronavirus which is taxonomically termed as SARS-CoV-2and named by World Health organization (WHO) as COVID-19 which emerged from Wuhan city, Hubei Province of China by the end of 2019 has caused unprecedented panic across the world. The rapid transmission of this virus from human to human made the World Health Organization (WHO) to declare this as the public health emergency of international concern and called it as global pandemic (Song & Karako, 2020). As on May 14, 2020, globally 4248389 COVID-19 cases have been reported and caused 292046deaths. Highest human casualty reported from USA with 109121 deaths (World Health Organization Situation Report, 2020).

The first case of the COVID-19 pandemic in India was reported on 30 January 2020. As on 17 May 2020, the Ministry of Health and Family Welfare, Government of India has reported 90927 confirmed cases from 33 states with 2872 deaths (MoHFW, 2020). Though India is in complete lockdown since March 24, over the weekend there is a rapid increase in COVID-19 cases in some states in India notably from Maharashtra, Gujarat, Tamil Nadu, Delhi, Madhya Pradesh and few other states. The rapid increase over the weekend in the month of May has created some kind of panic in India. The government and other civil bodies are making efforts to mitigate the spread of this virus.

Scientific literature on this pandemic is important in order to combat this novel coronavirus. Researchers across the world have been involved in identifying the cause, clinical features and developing possible vaccinations for COVID-19. As a result there is a rapid growth in publishing scholarly literature on the subject. Many academic institutions of international repute and also publication houses involving in scientific publishing industry have made their literature on COVID-19 or novel coronavirus freely available to the scientific community in particular for diagnosis, treatment and preventive strategies against the virus and public in general to create awareness of the infectious virus.

The Bibliometric studies which helps in quantifying the research publication pattern in a particular domain have also been done to assess the research productivity of scientific literature on COVID-19. Bibliometric studies help in identifying the emerging area of research, provide evidence of impact of research through citations, helps in identifying right scholarly literature to consult for study and also for carrying research forward, and also helpful for policy makers to strategize the potential research areas and funding. There were few bibliometric studies on COVID-19 publications pattern worldwide (Dehghanbanadaki et al, 2020; Hossain, 2020; Nasab, & Rahim, 2020). There is hardly any studies on country specific. In this regard, here an attempt has been made to look into the Indian contributions to COVID-19 research publications.

## 2. Data Source & Methods

The study used World Health organization (WHO) COVID-19 database made available through WHO official website under global research on coronavirus disease (COVID-19) as a data source for identifying the relevant literature on COVID-19. WHO COVID-19 database is curating global literature publishing on coronavirus since it declared COVID-19 a global pandemic. WHO says that the database represents a comprehensive multilingual source of current literature on the topic. This database extract information from various bibliographic databases, hand searching, and the addition of other expert-referred scientific articles and updates daily.

For current study the researcher used the following keywords to retrieve data on Indian Publication from WHO COVID-19 database. WHO has made its COVID-19 database searchable freely and data can be exported to .CSV and RIS format. The search terms used for retrieving the data were “COVID-19” and “India”. The “title, abstract and subject” option available on the database website was used to retrieve the documents. The search results retrieved107 results for the search term. The database was search on 12^th^ May 2020. The data was exported from .CSV format to excel sheet for further refinement of data and analysis. After thoroughly reviewing the data, it was found that there were few repeatable titles and titles not associated with Indian author or authors and articles other than English language were excluded from the study. Only articles written in English were included in the study. In all 89 articles were considered for the final analysis.

The study analyzed the date-wise Indian publication pattern on COVID-19, most prolific authors, institutions with highest publications, states with highest publication based corresponding authors state affiliation, top journals in which Indians have published their publications frequently and document type and research area, author keywords tree map analysis and highly cited COVID-19 documents of Indian authors. For retrieving the citation data for identifying the top cited documents, Google Scholar citation database was used. In order to determine the research focus or area of Indian publications on COVID-19, the author used the similar subject categories of earlier study done by Lou, et al (2020) on coronavirus. For identifying journal impact factor, the Scopus CiteScore and SCImago Journal Ranking & Country Ranking was consulted.

## 3. Data Analysis and Results

The study used descriptive statistics to analyze and visualized the data. The data analysis and result of the study is presented here in this section.

### 3.1. Date-Wise Publications Pattern

Figure-1 indicates the date-wise daily publication pattern of Indians on COVID-19. All the publications that have been included in the study were curated from March 03, 2020 to May 08, 2020 in WHO COVID-19 database. As it can be seen from Table-1 that after mid-April there is a considerable and constant increase in number of publications published from Indian authors on the subject. Given the gravity of the problem and the virus infectivity scale, it is obvious that medical researchers have been looking for potential treatment or vaccines for this virus and this has increased the output of research in India and in other parts of the globe as well. Clinical trials for identifying vaccine or potential drug for COVID-19 is in full swing. The clinicaltrials.gov database has listed 1510 studies on COVID-19. This indicates the increasing interest and rapidity in which research studies have been carried out on this virus.

**Figure-1:**
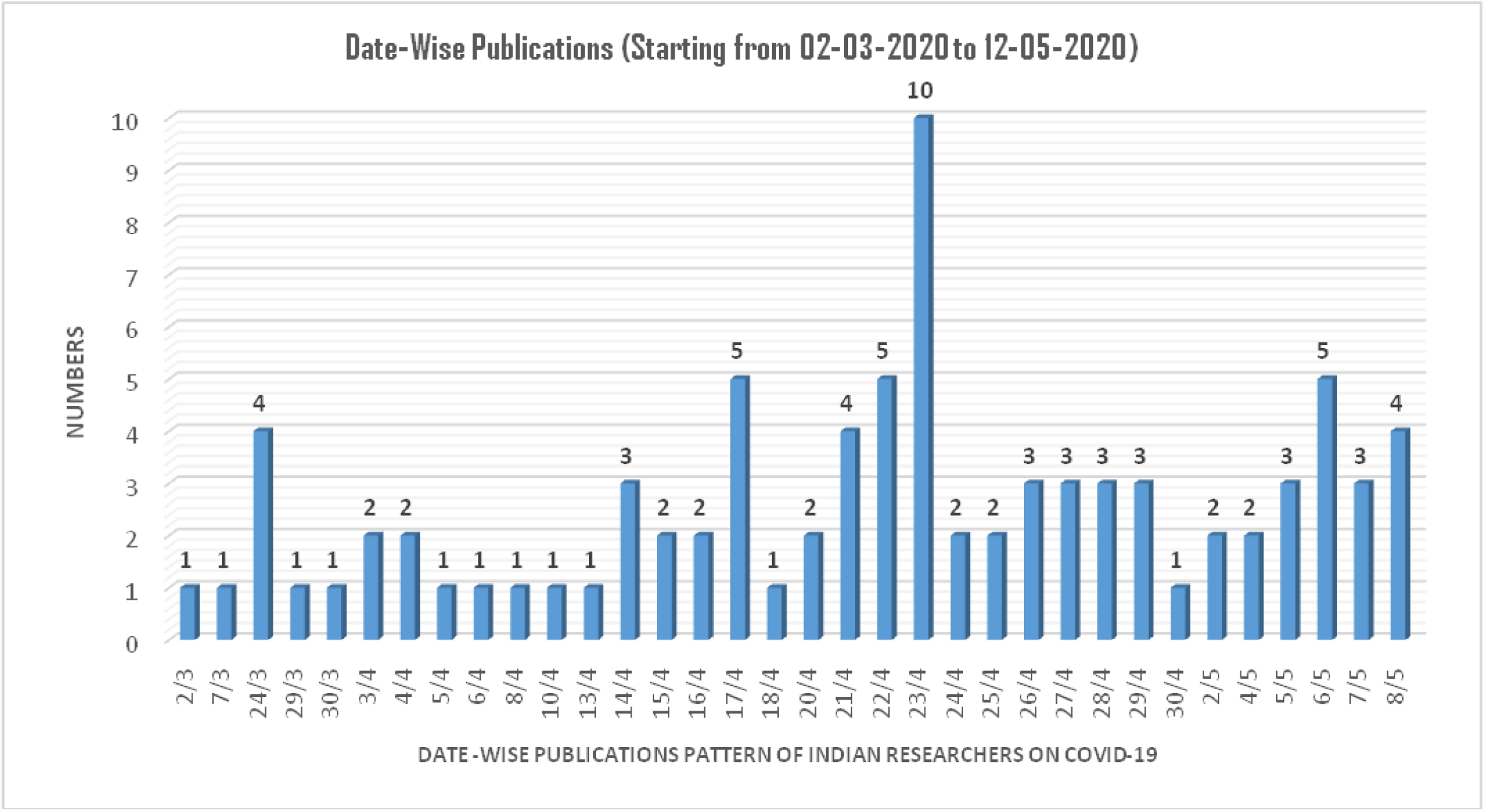
Date-Wise Indian Publications on COVID-19.

**Table-1:** Author(s) with Three or More Publication on COVID-19.

| **Author Name** | **Institutional Affiliations** | **Number of Publications** | **% of 89** |
| --- | --- | --- | --- |
| Gupta, Nivedita | Division of Epidemiology & Communicable Diseases, Indian Council of Medical Research, New Delhi, India | 06 | 6.74 |
| Bhatnagar, Tarun | National Institute of Epidemiology-ICMR, Chennai, Tamil Nadu, India | 04 | 4.49 |
| Gangakhedkar, Raman R. | Division of Epidemiology & Communicable Diseases, Indian Council of Medical Research, New Delhi, India | 04 | 4.49 |
| Giri, Sidhartha | Division of Epidemiology & Communicable Diseases, Indian Council of Medical Research, New Delhi, India | 04 | 4.49 |
| Misra, Anoop | Fortis CDOC Hospital for Diabetes and Allied Sciences, New Delhi, India | 04 | 4.49 |
| Potdar, Varsh | National Institute of Virology-ICMR, Pune, Maharashtra, India | 04 | 4.49 |
| Yadav, Pragya D. | National Institute of Virology-ICMR, Pune, Maharashtra, India | 04 | 4.49 |
| Abraham, Priya | Electron Microscopy & ICMR-National Institute of Virology, Pune, Maharashtra, India | 03 | 3.37 |
| Basu, Atanu | Maximum Containment Laboratory, ICMR-National Institute of Virology, Pune, Maharashtra, India. | 03 | 3.37 |
| Choudhary, Manohar Lal | Bioinformatics & Data Management Group, ICMR-National Institute of Virology, Pune, Maharashtra, India. | 03 | 3.37 |
| Murhekar, Manoj V. | National Institute of Epidemiology-ICMR, Chennai, Tamil Nadu, India. | 03 | 3.37 |
| Nyayanit, Dimpal A. | Maximum Containment Laboratory, ICMR-National Institute of Virology, Pune, Maharashtra, India. | 03 | 3.37 |
| Shete-Aich, Anita | Maximum Containment Laboratory, ICMR-National Institute of Virology, Pune, Maharashtra, India. | 03 | 3.37 |

### 3.2. Most Prolific Indian Authors on COVID-19 Research Publications

Table-1 depicts the most prolific Indian authors who have published three or more articles. Of the 89 articles, only 13 (14.60%) articles were single authored remaining articles were authored collaboratively. Nivedita Gupta of Division of Epidemiology & Communicable Diseases, Indian Council of Medical Research, New Delhi, topped the Table with 6 (6.74%) publications to her credit on COVID-19 research in India. Tarun Bhatnagar, National Institute of Epidemiology-ICMR, Chennai, Tamil Nadu, Raman R Gangakhedkar, SidharthaGiri of Division of Epidemiology & Communicable Diseases, Indian Council of Medical Research, New Delhi have authored 4 articles each. Other authors with three or more publications can be seen in Table-1. Anoop Misra is the only outlier in the Table. The other authors listed in the Table either have associated with Indian Council for Medical Research (ICMR), New Delhi or National Institute of Virology, Pune which is also part of ICMR. Except Anoop Misra of Fortis CDOC Hospital, New Delhi, the top published authors have published their articles together collaboratively. Nivedita Gupta, Bhatnagar, Gangakhedkar and Giri have published together and other group consisting of Potdar, Yadav, Abraham and other from NIV have published together.

### 3.3. State-Wise Publications Based on Corresponding Author’s State Affiliation

The state-wise publication profile was created by using corresponding author state affiliation. The geographic heat map (Figure-2) indicates state-wise publications on COVID-19. Delhi with 20 (22.47%) publications has produced highest number of publications on this subject, West Bengal with 11 (12.36%) publications second in the order followed by Maharashtra with 9 (10.11%) publications. Chandigarh, Karnataka and Tamil Nadu have 4 publications each with 4.49%. Haryana, Punjab and Rajasthan have published 3 (3.37%) papers each. In Northern part, Delhi, Punjab and smaller state like Chandigarh have performed better. BIMARU states like Madhya Pradesh, Uttar Pradesh have published only one article each, Bihar has not published even a single paper on this topic. These states are largest in terms of their population. No research publications on COVID-19 have appeared from North Eastern region. It is right time to increase the research output and have better healthcare facilities across India, instead situating in few pockets.

**Figure-2:**
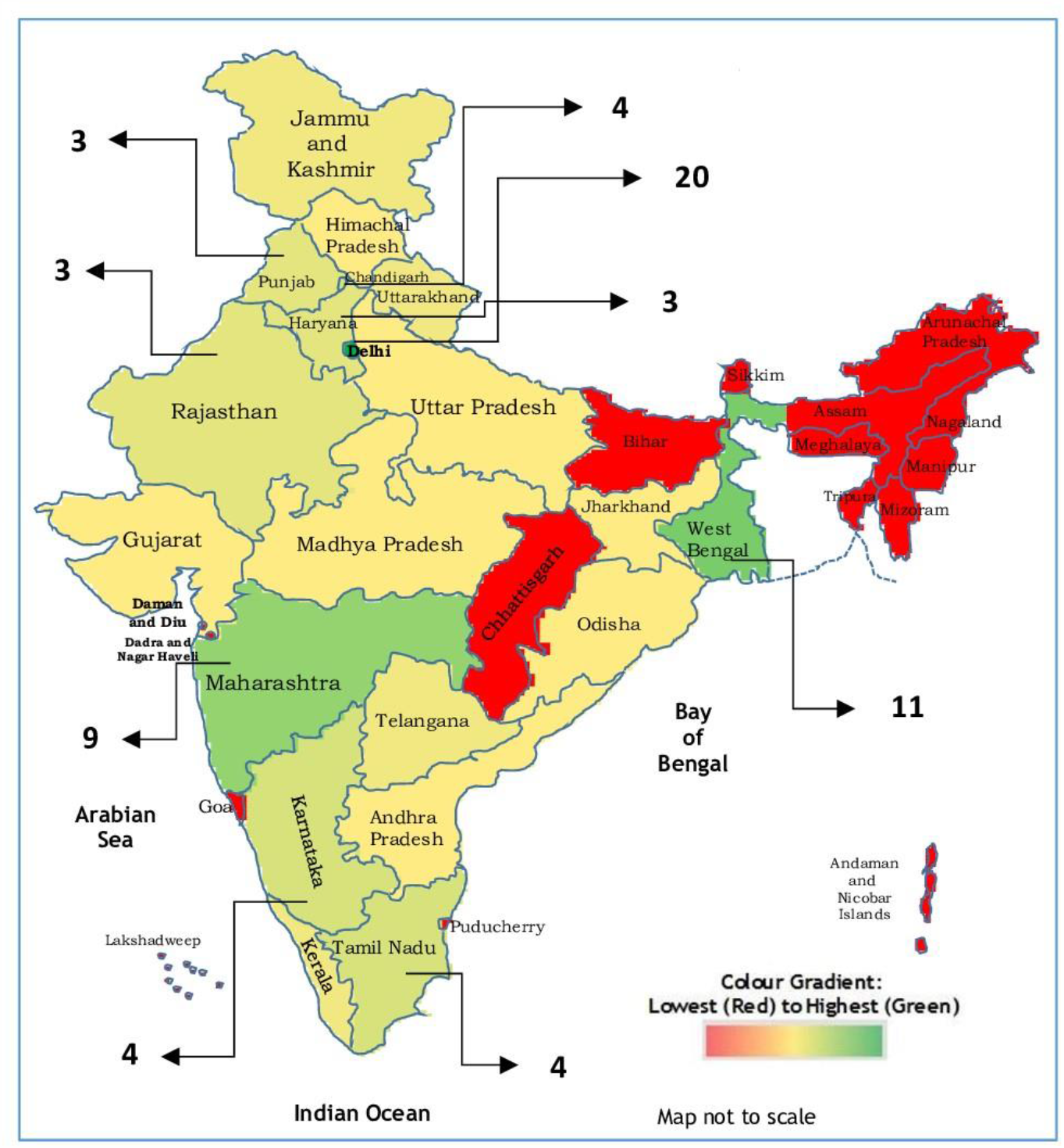
Indian State(s) Publications Based on the Correspondence Author’s Affiliation^1^.

### 3.4. Most Prolific Indian Institutions on COVID-19 Research

The bubble chart (Figure-3) depicts the Indian institutions which have published three or more publications on COVID-19 or coronavirus related works. Two hundred and forty two institutions have involved in publishing 89 papers, eight institutions have published three or more publications. The All India Institute of Medical Sciences (AIIMS), New Delhi with 8 (3.31%) publications is in the top positions followed by Post Graduate Institute of Medical Education & Research (PGIMER), Chandigarh with 7 publications (2.89%) and Indian Council of Medical Research (ICMR), New Delhi and National Institute of Virology, Pune with 6 (2.48%) publications each. National Institute of Epidemiology, Tamil Nadu and Fortis CDOC Hospital for Diabetes and Allied Sciences, New Delhi have 4 (1.65%) publications each. Armed Force Medical College, Pune and Maulana Azad Medical College, New Delhi have published 3(1.24%) papers each. It is notable that AIIMS and ICMR are two premier medical institutions in India and have been guiding Government of India in combating the lethal infectious disease COVID-19. It is worth mentioning that, the National Institute of Virology, Pune has recently developed IgG ELISA test kit for coronavirus for speeding the antibody test, which is a first of its kind in India (Prasad, 2020).

**Figure-3:**
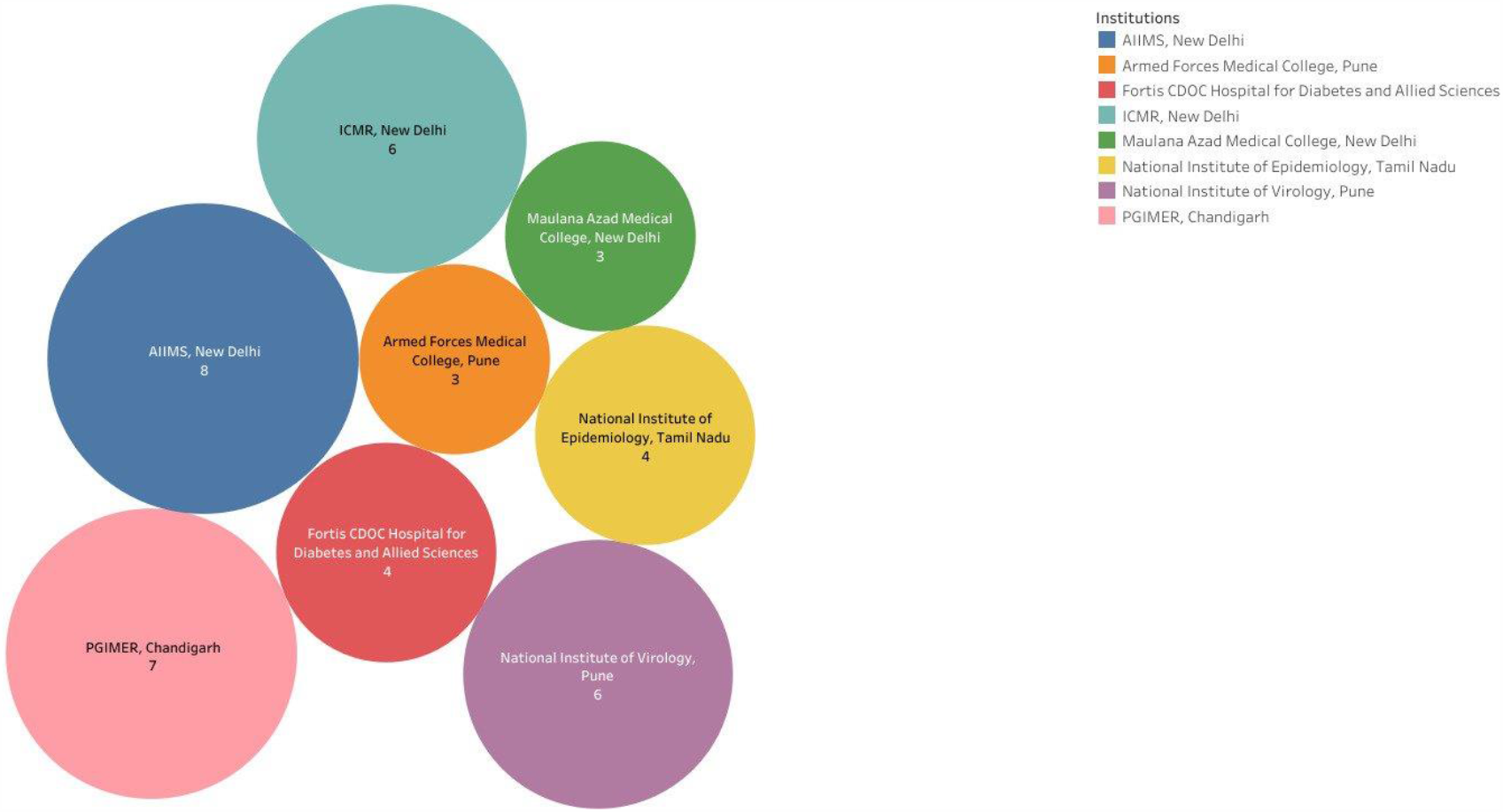
Most Prolific Institutions on COVID-19 Research through Bubble Chart.

### 3.5. Most Prolific Journals in Which Indian’s Have Published their Works on COVID-19 Frequently

Indians have published their publications on novel coronavirus or COVID-19 in 51 journals. Table-2 presents the top journal in which publications were frequently appeared. The Indian Journal of Medical Research has emerged as the top journals with 14 (15.73%) publications followed by Diabetes & Metabolic Syndrome: Clinical Research & Reviews and Science of the Total Environment with 6 (6.74%) publications each. Indian Journal of Ophthalmology has 4 (4.49%) publications to its credits and it’s in third position in the Table. International Journal of Research in Pharmaceutical Sciences and VirusDisease have 3 (3.37%) publications to their credits each and stands at fourth position. In all, these 13 journals have published 42 research papers, which accounts for 56.19% of the total publications. All the journals which are even under paywall have made their preprints available freely for researchers working on SARS-CoV-2 across the globe for speeding up for identifying potential vaccines or drugs for this virus.

**Table-2:** Top Journals in Which Indian COVID-19 Research Published Frequently.

| SL No. | Journal Name | Publisher | Number of Publications | % of 89 | Journal CiteScore* |
| --- | --- | --- | --- | --- | --- |
| 1. | Indian Journal of Medical Research | Wolters Kluwer - Medknow | 14 | 15.73 | 1.09 |
| 2. | Diabetes & Metabolic Syndrome: Clinical Research & Reviews | Elsevier BV | 06 | 6.74 | 2.08 |
| 3. | Science of the Total Environment | Elsevier BV | 06 | 6.74 | 5.92 |
| 4. | Indian Journal of Ophthalmology | Wolters Kluwer Health | 04 | 4.49 | 0.81 |
| 5. | International Journal of Research in Pharmaceutical Sciences | JK Welfare & Pharmascope Foundation | 03 | 3.37 | 0.21 |
| 6. | VirusDisease | Springer India | 03 | 3.37 | 0.97 |
| 7. | Asian Journal of Psychiatry | Elsevier BV | 02 | 2.25 | 1.29 |
| 8. | C&EN Global Enterprise | ACS Publications | 02 | 2.25 | - |
| 9. | Infection Control & Hospital Epidemiology | Cambridge University Press | 02 | 2.25 | 2.09 |
| 10. | Medical Journal Armed Forces India | Elsevier India | 02 | 2.25 | 0.55 |
| 11. | Monaldi Archives for Chest Disease | Page Press Publications | 02 | 2.25 | 0.53 |
| 12. | JMIR Public Health & Surveillance | JMIR Publications Inc | 02 | 2.25 | - |
| 13. | The Lancet | The Lancet Publishing Group | 02 | 2.25 | 10.28 |

\* Based on 2018 Scopus CiteScore metrics

### 3.6. Research Area and Document Type of COVID-19 Publications

Figure-4 indicates the research area and the document type of the Indian publications. Of the 89 publications 38 (42.70%) were articles, 13 (almost 15%) were of letters to editor or correspondence, 12 (13.48%) were commentary/opinion/perspective/viewpoints kind of documents, 7 (7.87%) were reviews, 6 (6.74%) were of guidelines/protocol/report related publications, another same number of documents were news items, 5 (5.62%) were editorials and 2 (2.25%) were short communications. If one were to see the document type, all kinds of documents ranging from articles to short communications were published. In terms of research area, of the 89 publications 75 of them were related to Epidemiology, which accounts for 84.27% of the total publications, 6 (6.74%) were related to Diagnosis & Treatment, 4 (4.49%) publications were on virology and 3 (3.37%) publications were of laboratory Examinations related studies and 1 publication was on clinical features of COVID-19 patients. Figur-4 clearly indicates the Indian research publication pattern with regard research areas. There were lot of studies on Epidemiology and its impact on diabetes, cardiologic patients and pandemic outbreak in India, less on clinical and laboratory examinations of COVID-19 patients, virology which are important for finding drugs or vaccine for the virus.

**Figure-4:**
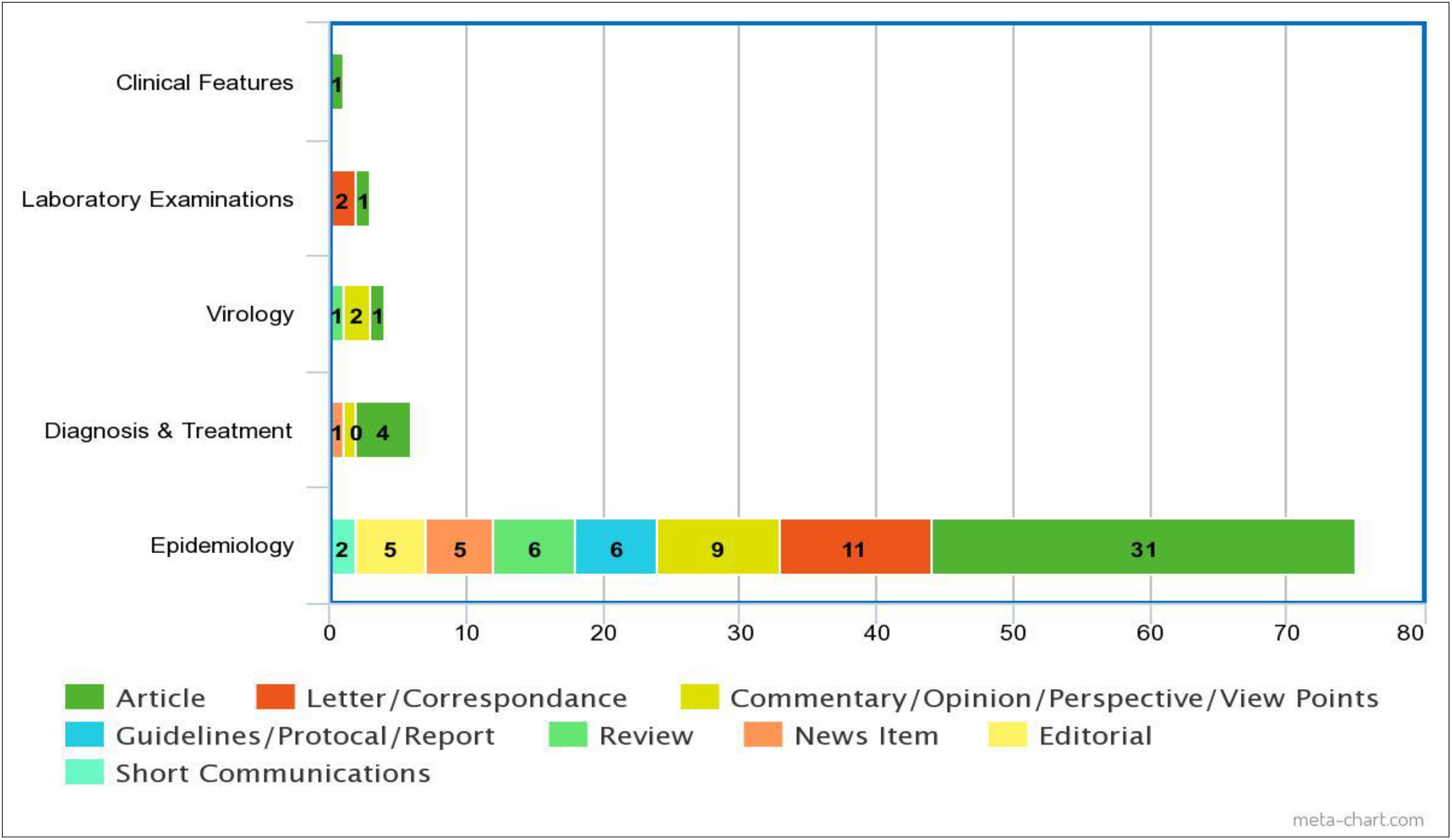
Research Area and Document Type on COVID-19.

### 3.7. COVID-19 Publications Keyword Frequency

The hierarchical tree map (Figure-5) indicates the frequency of author assigned keywords to the documents that they have published on COVID-19. Keywords are helpful in identifying key domains of research and its growth over a period of time. The keywords of corpus contained 415 words with 208 unique words, same have been used to identify the frequency of keywords on COVID-19. It is found that “covid” (34 times), “coronavirus” (23 times), “India” (14 times), “pandemic” (12 times), “sars” (8 times), “cov”, “health”, “management”, “syndrome” found 6 times each. Other keywords frequently appeared in the document can be seen in Figure-5.

**Figure-5:**
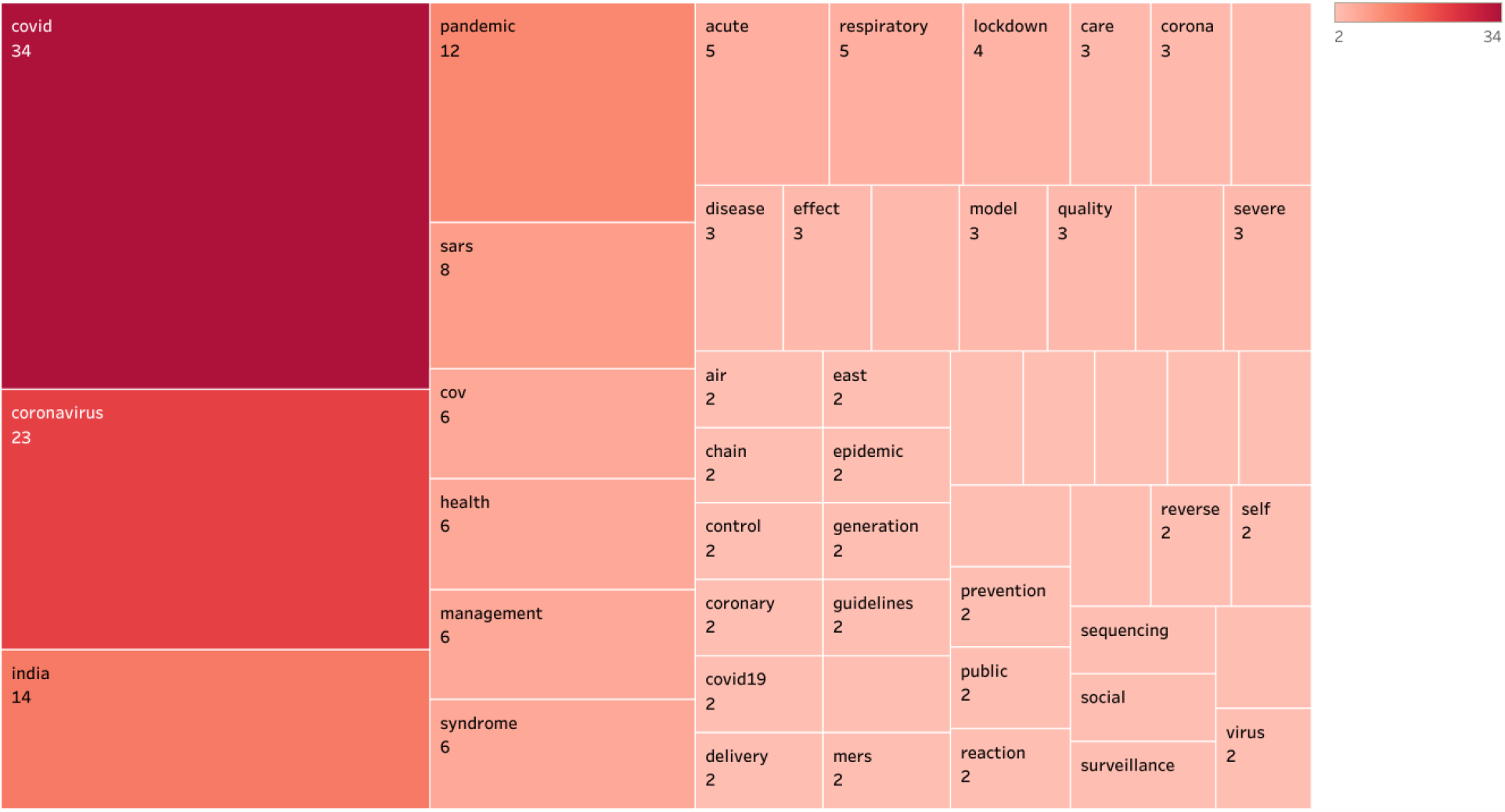
Hierarchical Tree Map of Frequency of Keywords on COVID-19.

### 3.8. Top Five Highly Cited Indian Publications on COVID-19

Eighty Nine articles have received 186 citations in all with an average citations of 2.18 per documents. Table-3 presents top five highly cited publications of Indian authors on novel coronavirus. The article entitled “Chloroquine and hydroxychloroquine in the treatment of COVID-19 with or without diabetes: A systematic search and a narrative review with a special reference to India and other developing countries” authored by Awadhesh Kumar Singh, et al and “Prudent public health intervention strategies to control the coronavirus disease 2019 transmission in India: A mathematical model-based approach” authored by Sandip Mandal et al, and “Fear of COVID 2019: First suicidal case in India!” by Kapil Goyal and others have received 28 citations each and topped the Table, followed by the article “Full-genome sequences of the first two SARS-CoV-2 viruses from India” authored by Pragya Yadav with 18 citations. Other highly cited articles can be seen Table-3.

**Table-3:**
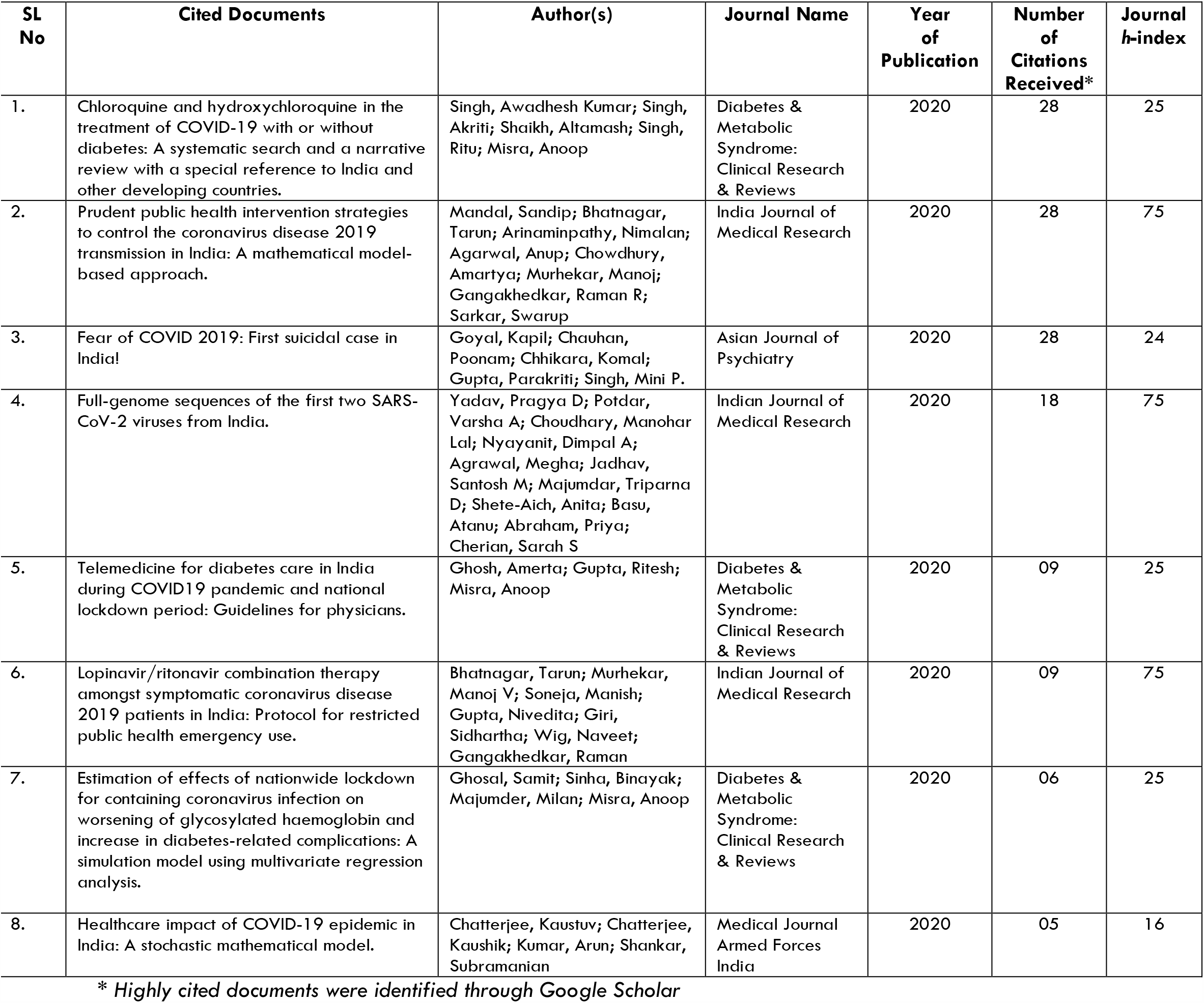
Top Five Highly Citd Indian Publications on COVID-19.

| SL No | Cited Documents | Author(s) | Journal Name | Year of Publication | Number of Citations Received* | Journal h-index |
| --- | --- | --- | --- | --- | --- | --- |
| 1. | Chloroquine and hydroxychloroquine in the treatment of COVID-19 with or without diabetes: A systematic search and a narrative review with a special reference to India and other developing countries. | Singh, Awadhesh Kumar; Singh, Akriti; Shaikh, Altamash; Singh, Ritu; Misra, Anoop | Diabetes & Metabolic Syndrome: Clinical Research & Reviews | 2020 | 28 | 25 |
| 2. | Prudent public health intervention strategies to control the coronavirus disease 2019 transmission in India: A mathematical model-based approach. | Mandal, Sandip; Bhatnagar, Tarun; Arinaminpathy, Nimalan; Agarwal, Anup; Chowdhury, Amartya; Murhekar, Manoj; Gangakhedkar, Raman R; Sarkar, Swarup | Indian Journal of Medical Research | 2020 | 28 | 75 |
| 3. | Fear of COVID 2019: First suicidal case in India! | Goyal, Kapil; Chauhan, Poonam; Chhikara, Komal; Gupta, Parakriti; Singh, Mini P. | Asian Journal of Psychiatry | 2020 | 28 | 24 |
| 4. | Full-genome sequences of the first two SARS-CoV-2 viruses from India. | Yadav, Pragya D; Potdar, Varsha A; Choudhary, Manohar Lal; Nyayanit, Dimpal A; Agrawal, Megha; Jadhav, Santosh M; Majumdar, Triparna D; Shete-Aich, Anita; Basu, Atanu; Abraham, Priya; Cherian, Sarah S | Indian Journal of Medical Research | 2020 | 18 | 75 |
| 5. | Telemedicine for diabetes care in India during COVID19 pandemic and national lockdown period: Guidelines for physicians. | Ghosh, Amerta; Gupta, Ritesh; Misra, Anoop | Diabetes & Metabolic Syndrome: Clinical Research & Reviews | 2020 | 09 | 25 |
| 6. | Lopinavir/ritonavir combination therapy amongst symptomatic coronavirus disease 2019 patients in India: Protocol for restricted public health emergency use. | Bhatnagar, Tarun; Murhekar, Manoj V; Soneja, Manish; Gupta, Nivedita; Giri, Sidhartha; Wig, Naveet; Gangakhedkar, Raman | Indian Journal of Medical Research | 2020 | 09 | 75 |
| 7. | Estimation of effects of nationwide lockdown for containing coronavirus infection on worsening of glycosylated haemoglobin and increase in diabetes-related complications: A simulation model using multivariate regression analysis. | Ghosal, Samit; Sinha, Binayak; Majumder, Milan; Misra, Anoop | Diabetes & Metabolic Syndrome: Clinical Research & Reviews | 2020 | 06 | 25 |
| 8. | Healthcare impact of COVID-19 epidemic in India: A stochastic mathematical model. | Chatterjee, Kaustuv; Chatterjee, Kaushik; Kumar, Arun; Shankar, Subramanian | Medical Journal Armed Forces India | 2020 | 05 | 16 |

\* Highly cited documents were identified through Google Scholar

If one were to simply observe the highly cited documents, it can be seen that publications which are of virology related, diagnosis and treatment or clinical features and simulations or mathematical related studies have received more citation than general studies on epidemiology or pandemic. Eight out of six highly cited publications were appeared in two journals, they are: Diabetes & Metabolic Syndrome: Clinical Research & Reviews and Indian Journal of Medical Research.

## 4. Discussion and Conclusion

The results of the study reflect the current Indian scholarly publications on COVID-19 or SARS-CoV-2. Result of this study found some significant insights. The authors who have published more number of publications have all come from either AIIMS or ICMR institutes, in this case National Institute of Virology, Pune. Most of these authors have collaborated each other in publishing research papers on COVID-19 and have come from same institutions. This emphasizes that there is need to collaborate with other medical research institutions outside the intra-institute collaborations. More than eighty five percent of the articles on coronavirus were collaborative publications. Since COVID-19 is a global pandemic, collaborative research is the need of the hour to mitigate this infectious disease. The state-wise publication pattern indicates that except smaller states like Delhi, Chandigarh, Punjab and Haryana there are no significant publications from North India’s largest states like Uttar Pradesh, Madhya Pradesh and Bihar. This can be attributed for lack of medical or health science education and research institutions in this part. This is high time that the Government of India establishes medical research institutes and strengthen healthcare facilities across India. There is no publications from North Eastern Region states also. This region is also in need of robust medical infrastructure.

In terms of institutional publications profile, again ICMR, PGIMER and AIIMS and National Institute of Virology, National Institute of Epidemiology and other few have three or more publications. One of the interesting fact is that it is the government medical research institutions that have published large number of publications on COVID-19. AIIMS and ICMR, New Delhi and its other institutions in other parts of India, like National Institute of Virology, Pune and National Institute of Epidemiology, Tamil Nadu have shared largest publications on COVID-19. China and USA are way ahead of India in terms of their research publications on COVID-19 compared to their GDP and publications per million populations (Chahrour et al, 2020). Central and State Universities which are in large numbers in India and they should accelerate their research capabilities at this time of global health crisis. In terms of journals, it is Indian Journal of Medical Research (IJMR) which has published more number of articles than any other journals, it accounts for almost 16% of total publications. ICMR and its other associated institutions have published largely on this journal on COVID-19.

Indian research is largely done in the area of epidemiology and its impact on diabetes, cardio patients, environmental impact and pandemic outbreak, less studies on clinical prognosis, pharmaceutical interventions, and laboratory based studies or virology related studies on SARS-CoV-2. The highly cited publications were of evidenced based studies, for instance “Chloroquine and hydroxychloroquine in the treatment of COVID-19 with or without diabetes: A systematic search and a narrative review with a special reference to India and other developing countries” or “Prudent public health intervention strategies to control the coronavirus disease 2019 transmission in India: A mathematical model-based approach” which have been cited by 28 times and other studies as well. As this pandemic is no way to go early, the research is shifting from basic to experimental studies across the globe (Nasab & Rahim, 2020), if we see the top highly cited documents in India also gradually studies are shifting more towards evidence based medical research for finding drugs or vaccine at the earliest for this highly infectious disease.

This bibliometric study though provides a bird-eye-view of the publications pattern of Indians on COVID-19, one must be a bit cautious in generalizing the results of this study. WHO COVID-19 database curated only expert-referred scientific articles and literature available through LitCOVID database of National Library of Medicine (NLM). It has not included the articles available through preprint servers or central subject repositories such as medRxiv and bioRxiv or arXiv e-print server o SSRN repository. It is suggested to take up a similar study using dimension.ai or semantic scholar’s CORD-19 dataset which includes articles deposited in preprint servers on COVID-19 to see the different publications pattern of Indian Publications on novel coronavirus or COVID-19 in a much larger scale. The dynamic nature of inclusion of literature to the WHO database on a daily basis is also be considered while generalizing this study.

## Data Availability

https://search.bvsalud.org/global-literature-on-novel-coronavirus-2019-ncov/

## Footnotes

1 *This geographic heat map is generated by using freely available excel template developed by www.indzara.com*

